# Diabetes mellitus, glycemic traits, and cerebrovascular disease: a Mendelian randomization study

**DOI:** 10.1101/2019.12.27.19015834

**Authors:** Marios K. Georgakis, Eric L Harshfield, Rainer Malik, Nora Franceschini, Claudia Langenberg, Nicholas J. Wareham, Hugh S. Markus, Martin Dichgans

## Abstract

**Rationale:** Type 2 diabetes mellitus (T2D) is an established risk factor for cerebrovascular disease but the mechanisms underlying this association remain elusive. Disentangling the causal effects of T2D, hyperglycemia, and pre-diabetic phenotypes (insulin resistance, β-cell dysfunction) on major etiological stroke subtypes (ischemic stroke, intracerebral hemorrhage, ischemic stroke subtypes) could inform the development of preventive strategies.

**Objective:** We employed Mendelian randomization (MR) to explore the effects of genetic predisposition to T2D, hyperglycemia, insulin resistance, and β-cell dysfunction on risk of stroke subtypes and related cerebrovascular phenotypes.

**Methods and Results:** We selected instruments for genetic predisposition to T2D, HbA1c levels, fasting glucose levels, insulin resistance, and β-cell dysfunction (proxied by pro-insulin levels) based on published genome-wide association studies (up to 898,130 individuals). Applying two-sample MR, we examined associations with ischemic stroke, intracerebral hemorrhage, and ischemic stroke subtypes (large artery, cardioembolic, small vessel stroke; up to 60,341 cases and 454,450 controls). We further explored associations with the related phenotypes of carotid atherosclerosis, imaging markers of cerebral white matter integrity, and brain atrophy. Genetic predisposition to T2D and elevated HbA1c levels in the pre-diabetic range were associated with higher risk of any ischemic stroke, large artery stroke, carotid plaque and small vessel stroke. Independently of HbA1c levels, we further found genetic predisposition to insulin resistance to be associated with large artery and small vessel stroke, whereas predisposition to β-cell dysfunction was associated with small vessel stroke. Predisposition to β-cell dysfunction was further associated with intracerebral hemorrhage, lower grey matter volume, and total brain volume.

**Conclusions:** This study supports causal effects of T2D and hyperglycemia on large artery and small vessel stroke. We show differential effects of genetically determined insulin resistance and β-cell dysfunction on large artery and small vessel stroke that might have implications for anti-diabetic treatments targeting these mechanisms.

## INTRODUCTION

Cerebrovascular disease is a major public health issue^1^ ranking as the second leading cause of mortality and adult disability worldwide.^2, 3^ Type 2 diabetes mellitus (T2D) is an established risk factor for cerebrovascular disease.^4, 5^ In cohort studies, T2D shows associations with higher risk for both ischemic and hemorrhagic stroke independently of other risk factors.^6^ Also, several studies found associations of measures of hyperglycemia (glycated hemoglobin (HbA1c) and fasting glucose levels) with risk of stroke, both in patients with and without diabetes.^6, 7^ However, large-scale randomized controlled trials (RCTs) testing intensive glucose-lowering in patients with T2D show no significant reductions in risk of stroke, possibly due to insufficient power.^8-11^ Moreover, the effects of T2D or hyperglycemia on etiological stroke subtypes (large artery stroke, cardioembolic stroke, small vessel stroke, intracerebral hemorrhage) remain elusive.

Currently available anti-diabetic medications act by either directly lowering glucose levels or by targeting two major mechanisms that contribute to hyperglycemia: insulin resistance or pancreatic β-cell dysfunction.^12, 13^ Observational data suggest that markers of insulin resistance, β-cell dysfunction, and hyperglycemia influence the risk of cardiovascular disease independently of each other.^14, 15^ However, data on stroke and its etiological subtypes are lacking. Moreover, there is a risk of confounding in observational studies. Developing targeted strategies for stroke prevention in patients at risk or suffering from T2D would require disentangling these relationships.

Mendelian randomization (MR) may help to clarify these associations. MR uses genetic variants as instruments for traits of interest and is not prone to confounding and reverse causation.^16^ As such, MR has proven to be a powerful methodology for inferring causality.^17, 18^ The availability of large-scale genome-wide association studies (GWAS) with detailed phenotyping of cases further enables the exploration of etiological stroke subtypes that are typically not considered in epidemiological studies.

Here, we leveraged data from large-scale GWASs and performed MR analyses, with the following aims: (i) to examine the effects of genetic predisposition to T2D on risk of ischemic stroke, ischemic stroke subtypes, and intracerebral hemorrhage; (ii) to explore the effects of genetically determined measures of hyperglycemia (HbA1c and fasting glucose levels) among diabetes-free individuals on these phenotypes; (iii) to disentangle the associations of genetic predisposition to insulin resistance, β-cell dysfunction, and hyperglycemia with major stroke etiologies; and (iv) to explore associations between diabetic traits and related vascular phenotypes including carotid atherosclerosis, neuroimaging markers of white mater integrity, and brain atrophy.

## METHODS

### Study design and data sources

This is a two-sample MR study following the guidelines for strengthening the reporting of Mendelian randomization studies (STROBE-MR).^19^ The data sources that we used are detailed in **Table 1**. The study is based on publicly available summary statistics from GWAS consortia. Our study design and the phenotypes explored in these analyses are depicted in **Figure 1**. We explored associations of genetic predisposition to T2D, measures of hyperglycemia (HbA1c and fasting glucose levels), as well as markers of insulin resistance and β-cell dysfunction with cerebrovascular disease phenotypes including stroke subtypes, carotid atherosclerosis, white matter (WM) integrity, and brain atrophy. Information on all genetic variants used as instruments in the current study are presented in **Supplementary Tables S1-S6**.

**Table 1.** Data sources that were used in the analyses for the current study.

| Phenotype | Source | N (Total or Cases/Controls) | Imputation reference panel | Ancestry | Adjustments |
| --- | --- | --- | --- | --- | --- |
| Diabetes mellitus type 2 | DIAGRAM Consortium<sup>20</sup> | 74,124/824,006 | HRC | European | age, sex, population structure |
| HbA1c | UK Biobank<sup>21</sup> | 408,989 | HRC + UK10K | White British | age, sex, population structure, genotyping platform array, assessment center |
| Fasting glucose levels | MAGIC Consortium<sup>22</sup> | 133,010 | HapMap | European | age, sex, population structure |
| Insulin resistance (fasting insulin levels) | Multi-trait GWAS<sup>25</sup> and MAGIC Consortium<sup>22</sup> | 108,557 | HapMap | European | age, sex, BMI, population structure |
| β-cell dysfunction (fasting proinsulin levels) | MAGIC Consortium<sup>26</sup> | 16,378 | 1000 Genomes | European | age, sex, fasting insulin, population structure |
| Any ischemic stroke | MEGASTROKE Consortium<sup>35</sup> | 60,341/454,450 | 1000 Genomes | Trans-ethnic (70% European) | age, sex, population structure |
| Large artery stroke | MEGASTROKE Consortium<sup>35</sup> | 6,688/454,450 | 1000 Genomes | Trans-ethnic (70% European) | age, sex, population structure |
| Cardioembolic stroke | MEGASTROKE Consortium<sup>35</sup> | 9,006/454,450 | 1000 Genomes | Trans-ethnic (70% European) | age, sex, population structure |
| Small vessel stroke | MEGASTROKE Consortium<sup>35</sup> | 11,710/454,450 | 1000 Genomes | Trans-ethnic (70% European) | age, sex, population structure |
| Intracerebral hemorrhage | ISGC meta-analysis<sup>38</sup> | 1,545/1,481 | 1000 Genomes | European | age, sex, population structure |
| Carotid plaque | CHARGE Consortium<sup>39</sup> | 21,540/26,894 | 1000 Genomes | European | age, sex, population structure |
| WMH volume | UK Biobank imaging database<sup>40</sup> | 17,534 | HRC + UK10K | White British | age, sex, mean resting and task functional MRI head motion, population structure, genotyping platform array |
| Mean diffusivity | UK Biobank imaging database<sup>40</sup> | 17,534 | HRC + UK10K | White British | age, sex, mean resting and task functional MRI head motion, population structure, genotyping platform array |
| Fractional anisotropy | UK Biobank imaging database<sup>40</sup> | 17,534 | HRC + UK10K | White British | age, sex, mean resting and task functional MRI head motion, population structure, genotyping platform array |
| Normalized grey matter volume | UK Biobank imaging database<sup>40</sup> | 17,534 | HRC + UK10K | White British | age, sex, mean resting and task functional MRI head motion, population structure, genotyping platform array |
| Normalized total brain volume | UK Biobank imaging database<sup>40</sup> | 17,534 | HRC + UK10K | White British | age, sex, mean resting and task functional MRI head motion, population structure, genotyping platform array |

**Figure 1.**
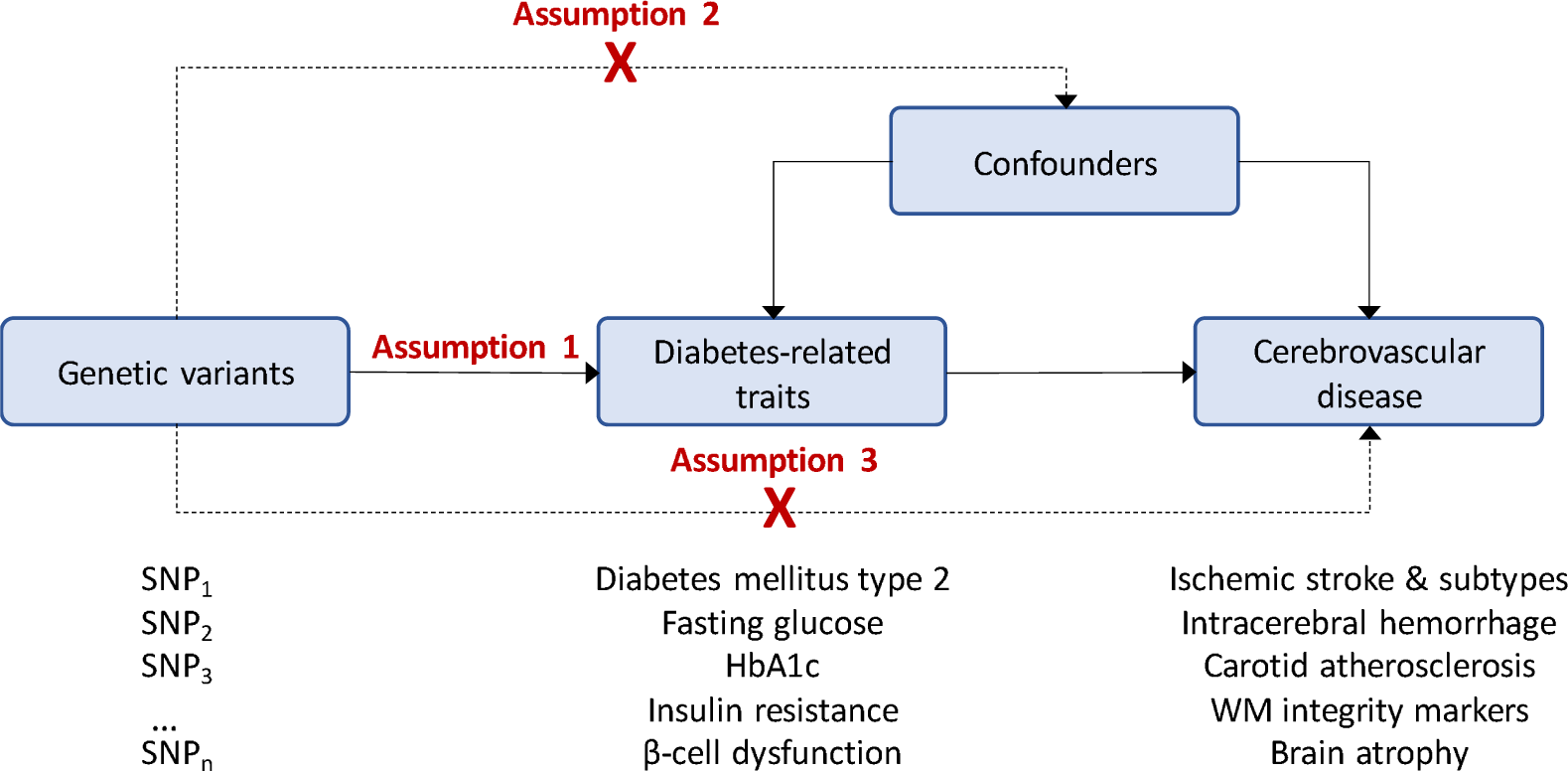
Schematic representation of our study design and assumptions of Mendelian randomization (MR) analyses. The assumptions of the MR study design include the following: the genetic variants (instruments) must be associated with the exposure (assumption 1); the variants must not be associated with confounders (assumption 2); the variants must influence the outcome only through the risk factor under study (assumption 3). *Abbreviations*. HbA1c, Glycated hemoglobin; SNP, single nucleotide polymorphism; WM, white matter.

### Genetic instrument selection

#### Diabetes mellitus type 2

We selected genetic instruments from the latest GWAS meta-analysis for T2D based on 74,124 cases and 824,006 controls of European ancestry from 32 studies included in the DIAGRAM consortium.^20^ The analyses were adjusted for age, sex, and population structure. We selected as genetic instruments all the 403 distinct genetic variants showing significant associations with T2D in this meta-analysis (either genome-wide significance at p<5×10^−8^ or independent locus-wide significance at p<10^−5^ at significant loci; **Supplementary Table S1**). In sensitivity analyses, we restricted our genetic instruments to those reaching genome-wide significance.

#### Hyperglycemia

We selected genetic instruments for HbA1c levels (per 1%-increment) based on a GWAS that we performed on 408,989 diabetes-free individuals of White British ancestry in the UK Biobank (UKB).^21^ We excluded individuals with self-reported history of physician-diagnosed diabetes, use of oral antidiabetic drugs or insulin, HbA1c level >6.5%, or random glucose levels >200 mg/dl. We adjusted for age, sex, genotyping platform array, assessment center, and the first 20 principal components of the population structure and performed the analyses using BOLT-LMM with correction for relatedness and subtle population stratification. For fasting glucose levels (per 1-SD increment), we used the most recent GWAS meta-analysis (adjusted only for age, sex, and population structure) by the MAGIC consortium on 133,010 diabetes-free individuals of European ancestry.^22^ For both HbA1c and fasting glucose, we selected as instruments genetic variants reaching genome-wide significance (p<5×10^−8^) after pruning for linkage disequilibrium at an r^2^<0.01 threshold. We identified 543 instruments for HbA1c levels and 21 for fasting glucose levels (**Supplementary Tables S2-S3**).^22^

As several genetic variants may influence HbA1c levels through effects on erythrocyte biology and not by inducing hyperglycemia,^23^ to isolate the effects of the hyperglycemia-related genetic component of HbA1c levels, we performed sensitivity analyses excluding those variants reported to be associated at p<0.001 with erythrocyte-related traits in Phenoscanner^24^ (hemoglobin concentration, red blood cell count, hematocrit, mean corpuscular volume, mean corpuscular hemoglobin concentration, mean corpuscular hemoglobin, red cell distribution width, reticulocyte count, reticulocyte fraction of red cells, immature fraction of reticulocytes, high light scatter percentage of red cells, high light scatter reticulocyte count).

#### Insulin resistance and β-cell dysfunction

As instruments for insulin resistance we used 53 genetic variants identified in a multi-trait GWAS to associate with all three components of this phenotype (fasting insulin levels, triglycerides and HDL-cholesterol; **Supplementary Table S4**).^25^ We weighted the instruments based on their effects on fasting insulin levels (per 1-log increment) in a GWAS meta-analysis of 108,557 diabetes-free European individuals.^22^ In accordance with existing literature, we proxied β-cell dysfunction based on fasting proinsulin levels (per 1 log-increment).^26-28^

We used summary statistics from a GWAS meta-analysis of 16,378 diabetes-free European individuals and identified 21 genetic instruments (at p<5×10^−8^ and r^2^<0.01; **Supplementary Table S5**).^26^ The GWAS for fasting insulin levels was adjusted for age, sex, and population structure,^22^ whereas the GWAS for pro-insulin was additionally adjusted for fasting insulin levels.^26^

We further used T2D-associated genetic variants previously grouped into clusters of diabetic endophenotypes; three clusters of insulin resistance (related to obesity, fat distribution, or lipid metabolism) and two clusters of β-cell dysfunction both associated with reduced levels of fasting insulin, but with opposing effects on fasting proinsulin.^29^ We used the clusters of the variants and the respective weights per variant and cluster, as described in the study by Udler *et al*. (**Supplementary Table S6)**. ^30^

### Proportion of explained variance and instrument strength

For all genetic variants used as instruments, we estimated the proportion of explained variance for the respective phenotypes (**Supplementary Tables S1-S5**). We estimated the variance explained by each genetic variant for T2D based on the method by So *et al*. for binary phenotypes^31^ and for the continuous traits we used a previously described formula based on summary statistics.^32^ For the estimations regarding T2D, we used a prevalence rate of 8.5%, according to the 2015 estimate of the global prevalence of the disease by the International Diabetes Federation.^33^ We then calculated the F-statistic as an indicator of instrument strength.^34^ In sensitivity analyses, we restricted our selection to variants with F>10, which is widely considered to indicate instruments with very low probability for weak instrument bias.^34^

### Associations with outcomes

We then examined associations of the selected instruments with ischemic stroke, ischemic stroke subtypes, and intracerebral hemorrhage (ICH) as the primary outcomes of interest. For ischemic stroke, we used summary GWAS data from the MEGASTROKE trans-ethnic population, mainly consisting of European individuals (70%).^35, 36^ We extracted summary GWAS statistics for any ischemic stroke (60,341 cases, 451,210 controls) and for the major ischemic stroke subtypes according to the TOAST (Trial of Org 10172 in Acute Stroke Treatment) classification:^37^ large artery stroke (6,688 cases, 238,513 controls), cardioembolic stroke (9,006 cases, 352,852 controls), and small vessel stroke (11,710 cases, 287,067 controls). GWAS data for ICH were derived from the International Stroke Genetics Consortium (ISGC) GWAS meta-analysis including 1,545 cases and 1,481 controls of European ancestry.^38^

Presence of carotid plaque, markers of WM tract integrity (WM hyperintensities (WMH) volume, mean diffusivity, fractional anisotropy), and markers of brain atrophy (grey matter volume, total brain volume) were explored as secondary outcomes. Carotid plaque data were derived from a GWAS meta-analysis (21,540 cases, 26,894 controls of European ancestry) from the CHARGE consortium.^39^ For the imaging phenotypes (WMH volume, mean diffusivity, fractional anisotropy, grey matter volume, total brain volume), we undertook GWAS analyses in the UK Biobank neuroimaging dataset including 17,534 individuals of White British ancestry based on the MRI sequences, as has been previously described.^40^ We performed linear regression analyses (additive models) for ln-transformed WMH volume, the first principal components of all measurements of mean diffusivity and fractional anisotropy across the different white matter tracts in the diffusion sequences, and for normalized grey matter and total brain volumes. Adjustments were made for age, sex, mean resting and task functional MRI head motion, the genotype platform array, and the first 10 principal components of the population structure.

### Statistical analysis

All analyses were performed in R (v3.5.0; The R Foundation for Statistical Computing) using the MendelianRandomization, TwoSampleMR, and the MR-PRESSO packages.

#### Main analyses

We applied two-sample MR using association estimates derived from the abovementioned sources. Following extraction of the SNP-specific association estimates between the instruments and the outcomes, and harmonization of the direction of estimates by effect alleles, we computed MR estimates for each instrument with the Wald estimator.^41^ We calculated standard errors with the Delta method. We then pooled individual MR estimates using random-effects inverse-variance weighted (IVW) meta-analyses.^41^ For the main analyses, we corrected for multiple comparisons with the false discovery rate (FDR) approach and set statistical significance at q-value<0.05. Associations not reaching this threshold, but showing an unadjusted p<0.05 were considered of nominal significance.

#### Assessment of pleiotropy and sensitivity analyses

MR estimates derived from the IVW approach could be biased in the presence of directional horizontal pleiotropy. As a measure of overall pleiotropy, we assessed heterogeneity across the SNP-specific MR estimates in the IVW MR analyses with the Cochran’s Q statistic (statistical significance set at p<0.05).^42^ We further applied alternative MR methods which are more robust to pleiotropic variants. The weighted median estimator allows the use of invalid instruments as long as at least half of the instruments used in the MR analysis are valid.^43^ The MR-Egger regression allows for the estimation of an intercept term that can be used as an indicator of unbalanced directional pleiotropy.^44^ MR-Egger provides less precise estimates and relies on the assumption that the strengths of potential pleiotropic instruments are independent of their direct associations with the outcome.^44^ The intercept obtained from MR-Egger regression was used as a measure of unbalanced pleiotropy (p<0.05 indicated significance).^44^ Finally, MR-PRESSO regresses the SNP-outcome estimates against the SNP-exposure estimates to test for outlier SNPs.^45^ Outliers are detected by sequentially removing all variants from the analyses and comparing the residual sum of squares as a global measure of heterogeneity (p<0.05 for detecting outliers); outliers are then removed and outlier-corrected estimates are provided.^45^ MR-PRESSO still relies on the assumption that at least half of the variants are valid instruments.^45^

#### Mediation analyses and multivariable Mendelian randomization

We additionally performed MR-based network mediation analyses^46^ to examine the extent to which increases in HbA1c levels mediated the effects of insulin resistance and β-cell dysfunction on stroke subtypes. We performed this analysis only for large artery and small vessel stroke, for which we found robust evidence of associations with HbA1c. We calculated the total effects of insulin resistance and β-cell dysfunction on the outcomes of interest using random-effects IVW MR analyses. For the direct effects, we combined the instruments for insulin resistance or β-cell dysfunction with those for HbA1c levels and performed multivariable MR analyses for their associations with the stroke subtypes.^47^ To calculate the indirect effects, we calculated the difference between the direct and the total effect estimates.^48^ We finally estimated the proportion of the overall effect of insulin resistance or β-cell dysfunction on stroke subtypes that was mediated through increases in HbA1C by dividing the indirect by the total effects.^48^ Standard errors were derived using the delta method.

## RESULTS

The 403 genetic variants used as genetic instruments for T2D explained 17.2% of the variance in T2D prevalence (**Supplementary Table S1**), whereas variants used as instruments for the continuous hyperglycemia traits, insulin resistance (proxied by fasting insulin levels), and β-cell dysfunction (proxied by fasting proinsulin), explained lower proportions of variance: 1.9% for HbA1c, 1.5% for fasting glucose, 0.7% for insulin resistance, and 4.5% for β-cell dysfunction (**Supplementary Tables S1-S5**).

### Genetic predisposition to type 2 diabetes mellitus and risk of stroke

In the primary IVW MR analyses, genetic predisposition to T2D (1-log-increment=2.72-fold higher odds) was significantly associated with a higher risk of any ischemic stroke (OR: 1.10, 95%CI: 1.07-1.12), large artery stroke (OR: 1.19, 95%CI: 1.14-1.26), and small vessel stroke (OR: 1.14, 95%CI: 1.10-1.19; **Figure 2**). In addition, there was an association of nominal significance with higher risk of cardioembolic stroke (OR: 1.05, 95%CI: 1.01-1.09). All SNPs used as insturments for genetic predisposition to T2D had F-statitics >10, thus indicating a very low probability of weak instrument bias. In sensitivity analyses restricted to SNPs reaching genome-wide significance (p<5×10^−8^) for association with T2D the results remained stable and statistically significant and we further found an association of nominal significance with ICH (**Supplementary Table S7**).

**Figure 2.**
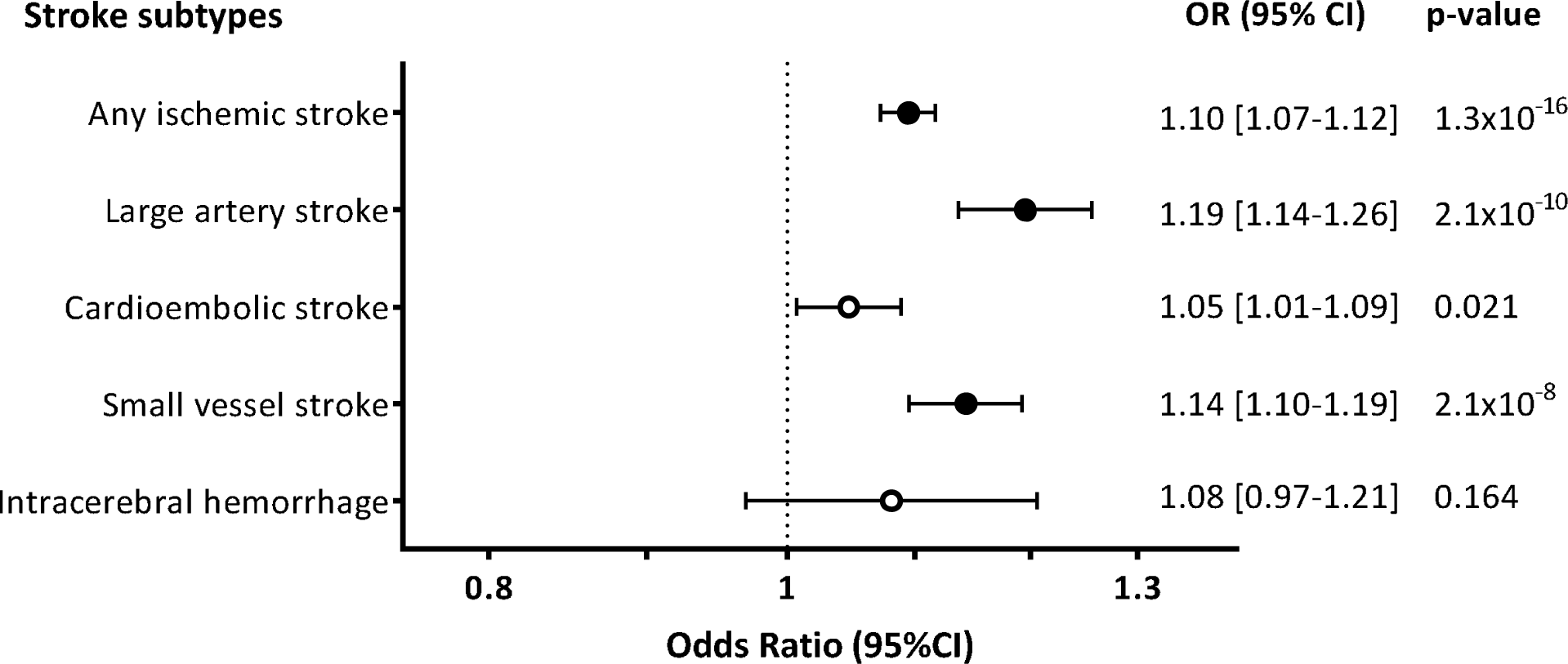
Mendelian Randomization associations between genetic predisposition to type 2 diabetes mellitus and stroke subtypes. Results derived from random-effects inverse-variance weighted analyses. Full circles correspond to statistically significant association estimates at an FDR-adjusted p-value<0.05.

With the exception of ICH, there was evidence of significant heterogeneity in all of the main analyses (p<0.05; **Supplementary Table S8**), but no evidence of unbalanced pleiotropy, as assessed by the Egger intercept p-values (all p>0.05; **Supplementary Table S7**). Across sensitivity analyses based on alternative MR methods (weighted median, MR-Egger, outlier-corrected MR-PRESSO), all effects remained directionally consistent and all estimates stable with p<0.05 for any ischemic stroke, large artery stroke, and small vessel stroke (**Supplementary Table S7**).

### Genetic predisposition to measures of hyperglycemia and risk of stroke

In analyses of hyperglycemia traits we found that genetically determined HbA1c levels (per 1%-increment) among diabetes-free individuals were significantly associated with risk of any ischemic stroke (OR: 1.28, 95%CI: 1.12-1.46), large artery stroke (OR: 1.61, 95%CI: 1.22-2.13), and small vessel stroke (OR: 1.83, 95%CI: 1.45-1.33; **Figure 3**). In contrast, we found no significant associations between genetically determined fasting glucose levels and risk of stroke subtypes (**Figure 3**).

**Figure 3.**
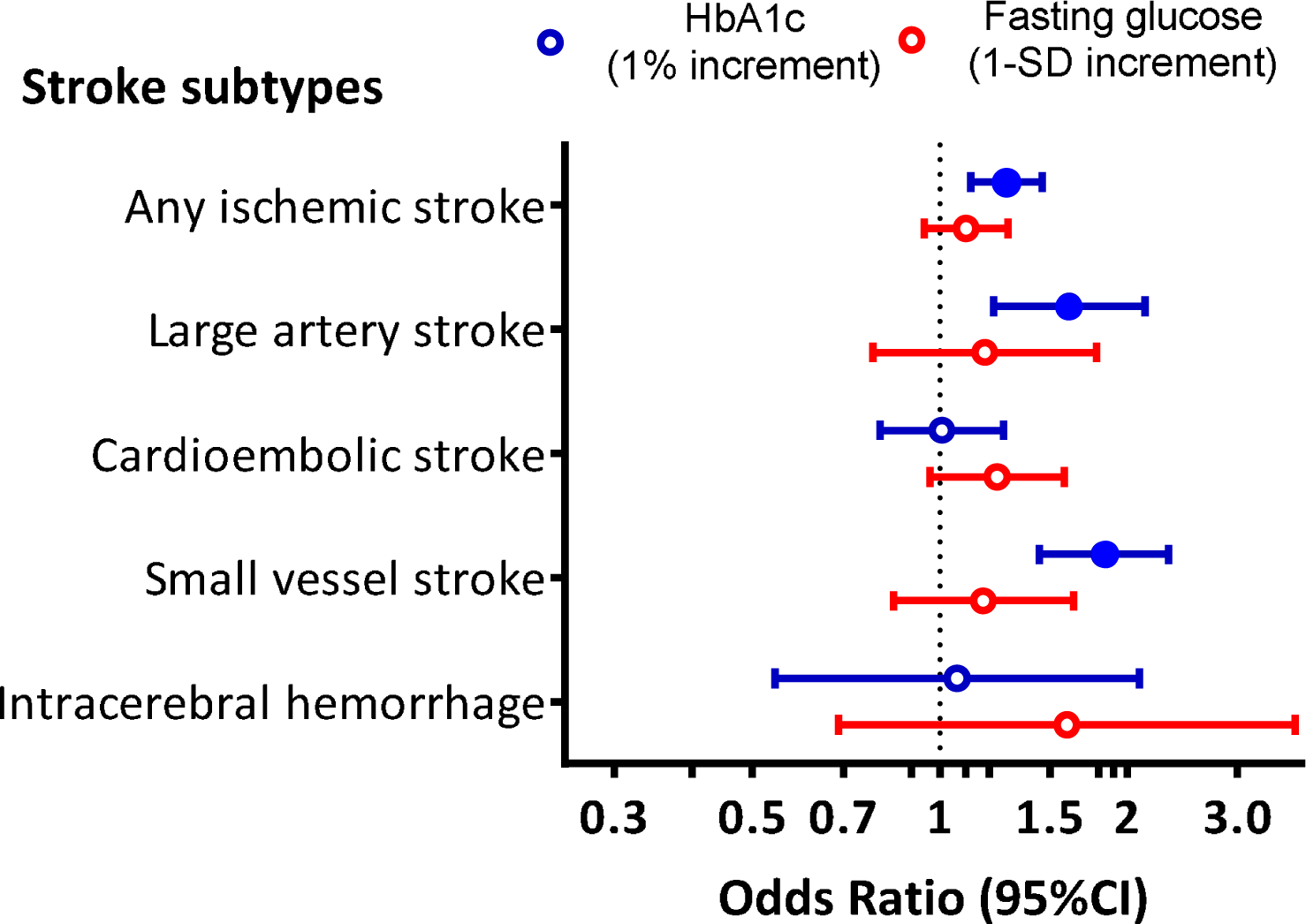
Mendelian Randomization associations of genetically determined hyperglycemia traits (HbA1c and fasting glucose) with stroke subtypes. Results derived from random-effects inverse-variance weighted analyses. Full colored circles correspond to statistically significant association estimates at an FDR-adjusted p-value<0.05. *Abbreviations*. HbA1c, Glycated hemoglobin.

There was evidence of heterogeneity in the analyses for HbA1c levels (**Supplementary Table S8**) and in MR-Egger the effect estimates for any ischemic stroke, large artery stroke, and small vessel stroke were smaller (**Supplementary Table S7**). However, in sensitivity analyses that excluded SNPs influencing HbA1c levels through erythrocyte-related traits, the association estimates were even larger (ischemic stroke, OR: 1.60, 95%CI: 1.30-1.98; large artery stroke, OR: 2.05, 95%CI: 1.29-3.26; small vessel stroke, OR: 2.44, 95%CI: 1.64-3.65) and there was no evidence of heterogeneity (all p>0.10). Similarly, the results remained stable, when excluding SNPs with an F-statistic <10 that might introduce weak instrument bias in the analyses.

### Genetic predisposition to insulin resistance, β**-cell dysfunction, and risk of stroke**

We next selected genetic variants as instruments for insulin resistance and β-cell dysfunction, the two primary underlying mechanisms contributing to the development of hyperglycemia and T2D. Among diabetes-free individuals, we found genetic predisposition to insulin resistance (1-log increment in fasting insulin levels) to be associated with a higher risk for ischemic stroke (OR: 1.33, 95%CI: 1.13-1.57), large artery stroke (OR: 1.60, 95%CI: 1.12-2.31), and small vessel stroke (OR: 1.63, 95%CI: 1.21-2.20; **Figure 4A**). Genetic predisposition to β-cell dysfunction (1-log increment in fasting proinsulin levels) was further associated with a higher risk for both small vessel stroke (OR: 1.38, 95%CI: 1.17-1.63) and ICH (OR: 1.75, 95%CI: 1.21-2.52). There was no heterogeneity in these analyses (**Supplementary Table S8**) and the results were consistent in alternative MR analyses (**Supplementary Table S7**).

**Figure 4.**
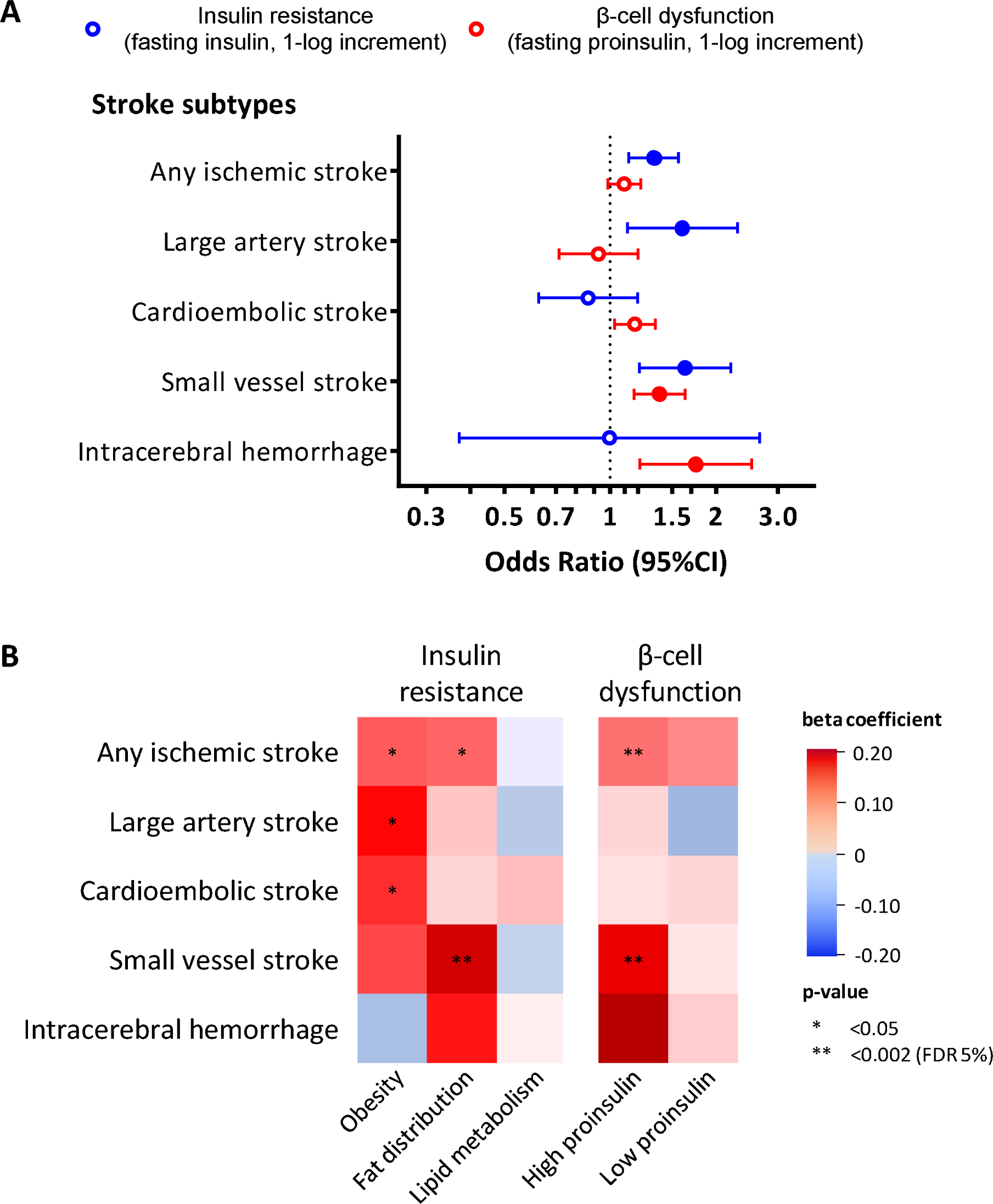
Mendelian Randomization associations of genetically determined insulin resistance and β-cell dysfunction with stroke subtypes. **(A)** Results derived from random-effects inverse-variance weighted analyses. (B) Heatmap of the associations between clusters of diabetic endophenotypes related to β-cell dysfunction and insulin resistance with the risk of stroke subtypes. Full colored circles in panel A correspond to statistically significant association estimates at an FDR-adjusted pvalue < 0.05.

To gain additional insights in the relationship between insulin resistance, β-cell dysfunction, and etiological stroke subtypes, we further explored the effects of T2D-associated variants clustered in five different mechanisms of action. These included three clusters for insulin resistance (mediated by obesity, fat distribution, lipid metabolism) and two clusters related to β-cell dysfunction (associated with high or low proinsulin). In multivariable analyses including all clusters and also adjusting for their effects on HbA1c, we found significant effects of genetic predisposition to β-cell dysfunction related to high proinsulin on risk of ischemic stroke and small vessel stroke (**Figure 4B**). We further found genetic predisposition to insulin resistance mediated through altered fat distribution to be associated with higher risk of small vessel stroke. Genetic predisposition to insulin resistance mediated through obesity showed associations of nominal significance with large artery and cardioembolic stroke.

We next explored whether the effects of genetic predisposition to insulin resistance and β-cell dysfunction on large artery and small vessel stroke are mediated through hyperglycemia (**Figure 5**). Genetic predisposition to insulin resistance was associated with higher risk of large artery stroke independently of its effects on HbA1c; the indirect effect through increases in HbA1c levels was not significant. For small vessel stroke, we found genetic predisposition to both insulin resistance and β-cell dysfunction to contribute to a higher risk independently of HbA1c levels; again, the indirect effects through HbA1c were not significant. HbA1c was associated with a higher risk of large artery and small vessel stroke, independently of insulin resistance or β-cell dysfunction (**Figure 5**).

**Figure 5.**
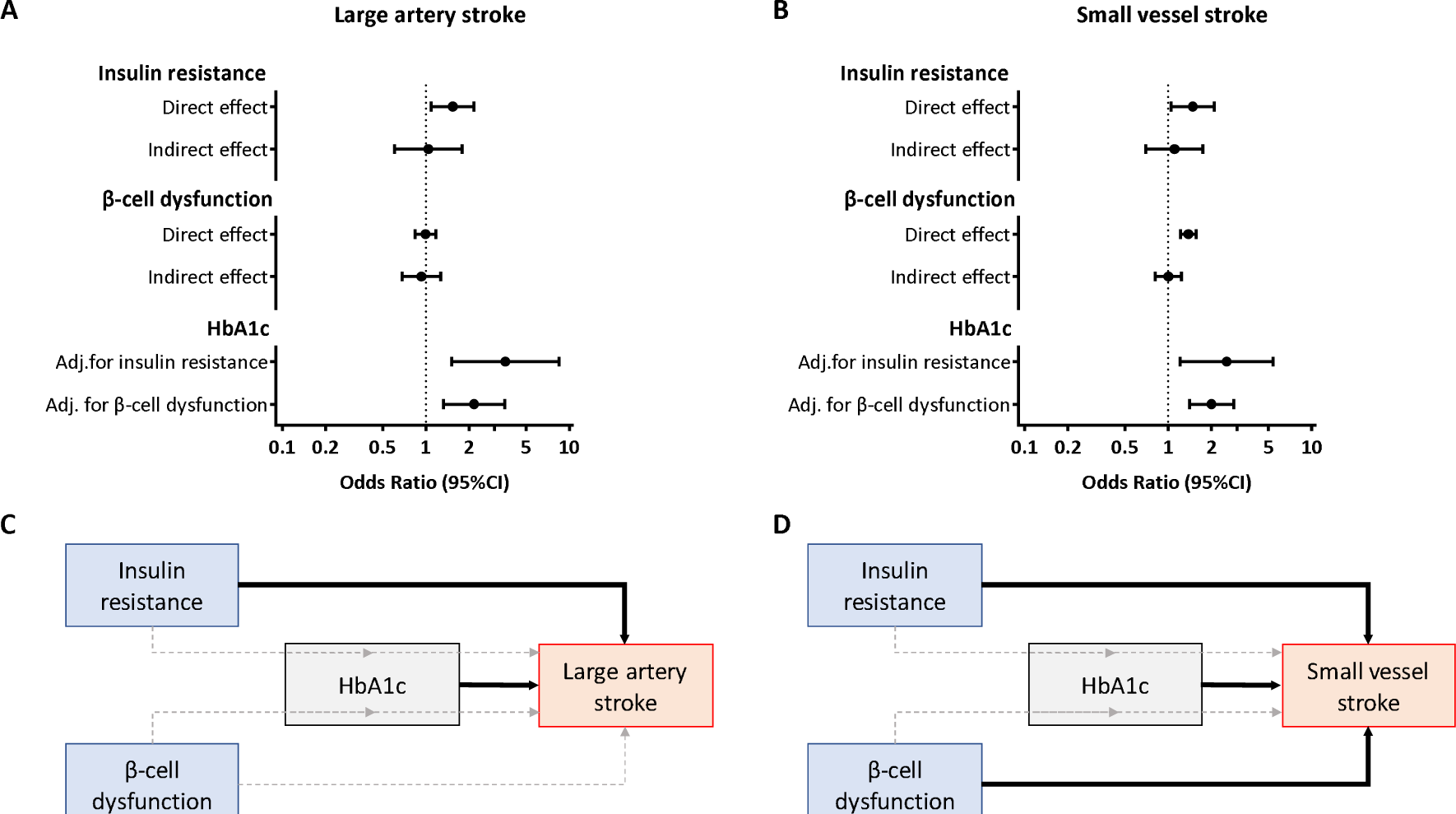
Mendelian randomization (MR)-based mediation analyses between genetically predicted insulin resistance, β-cell dysfunction, HbA1c levels and risk of large artery and small vessel stroke. (A, B) Direct and indirect (through HbA1c levels) effects of insulin resistance and β-cell dysfunction and effects of HbA1c adjusted for these phenotypes (by multivariable MR) on risk of large artery and small vessel stroke. (C, B) Schematic representation of the results. Dark arrows represent significant associations, whereas light grey dotted lines represent non-significant associations. *Abbreviations*. HbA1c, Glycated hemoglobin A1c.

### Genetic predisposition to type 2 diabetes and glycemis traits and associations with etiologically related cerebrovascular phenotypes

**Table 2** presents the MR associations of genetic predisposition to T2D, measures of hyperglycemia, insulin resistance, and β-cell dysfunction, with carotid plaque, as well as with neuroimaging traits related to white matter integrity and brain atrophy. Genetic predisposition to T2D and genetically elevated HbA1c levels were associated with carotid plaque. We further found a significant association between genetic predisposition to T2D and lower fractional anisotropy, a diffusion imaging marker of impaired white matter tract integrity, as well as nominally significant associations with lower grey matter and total brain volumes (**Table 2**). Genetic predisposition to β-cell dysfunction (1-log increment in fasting proinsulin levels) was further associated with lower grey matter volume (beta: −0.13, 95%CI: −0.20 to −0.07) and total brain volume (beta: −0.17, 95%CI: −0.23 to −0.11; **Table 2**). These results remained stable in sensitivity analyses (**Supplementary Table S7**).

**Table 2.** Mendelian randomization associations between genetically determined diabetic traits and etiologically related cerebrovascular phenotypes, as derived from random-effects inverse-variance weighted analyses.

| Exposure | Carotid plaque<br>OR (95%CI) | White matter integrity |  |  | Brain atrophy |  |
| --- | --- | --- | --- | --- | --- | --- |
|  |  | WMH volume | Mean diffusivity | Fractional anisotropy | Grey matter volume | Total brain volume |
|  |  | beta (95%CI) |  |  |  |  |
| Diabetes mellitus type 2 | **1.08 (1.04-1.12)** | 0.003 (-0.016, 0.023) | 0.011 (-0.011, 0.033) | **-0.028 (-0.048, -0.006)** | -0.023 (-0.041, -0.005)<sup>a</sup> | -0.021 (-0.041, -0.001)<sup>a</sup> |
| HbA1c (1%-increment) | **1.52 (1.24-1.85)** | 0.047 (-0.071, 0.164) | 0.002 (-0.129, 0.133) | -0.091 (-0.206, 0.025) | -0.122 (-0.230, -0.014)<sup>a</sup> | -0.083 (-0.204, 0.039) |
| Fasting glucose levels (1 SD increment) | 1.16 (0.93-1.44) | -0.012 (-0.135, 0.111) | -0.060 (-0.175, 0.055) | -0.034 (-0.158, 0.090) | -0.072 (-0.191, 0.048) | -0.092 (-0.212, 0.027) |
| Insulin resistance (1 log-increment in fasting insulin levels) | 0.93 (0.83-1.05) | 0.094 (-0.062, 0.251) | 0.146 (-0.056, 0.347) | -0.181 (-0.380, 0.019) | -0.039 (-0.220, 0.142) | -0.087 (-0.285, 0.112) |
| β-cell dysfunction (1 log-increment in fasting proinsulin levels) | 1.10 (0.80-1.50) | 0.062 (-0.021, 0.146) | 0.048 (-0.017, 0.114) | -0.048 (-0.115, 0.020) | **-0.130 (-0.195, -0.065)** | **-0.170 (-0.232, -0.108)** |

Odds Ratios (OR) are presented for binary traits (carotid plaque) and beta coefficients (standardized based on the SD of the measure) for the continuous imaging traits.
**Bold** indicates statistical significance at an FDR-adjusted p-value<0.05.
<sup>a</sup> Associations reaching nominal significance (unadjusted p<0.05)

## DISCUSSION

Levaraging large-scale GWAS data in MR analyses, we investigated the causal associations between T2D, glycemic traits, and cerebrovascular disease. We found genetic predisposition to T2D and hyperglycemia (elevated HbA1c levels) to be associated with a higher risk of ischemic stroke, particularly large artery and small vessel stroke. Independently of hyperglycemia, genetic predisposition to insulin resistance but not β-cell dysfunction was associated with higher risk of large artery stroke, whereas genetic predisposition to both insulin resistance and β-cell dysfunction was associated with small vessel stroke. Genetic determinats for T2D and hyperglycemia further showed significant effects on carotid plaque and fractional anisotropy, a WM neuroimaging marker related to cerebral small vessel disease. Conversely, genetic predisposition to β-cell dysfunction was associated with intracerebral hemorrhage and neuroimaging markers of brain atrophy.

Our MR results provide genetic evidence for a causal effect of T2D, and also hyperglycemia in the non-diabetic range, on risk of ischemic stroke. While T2D is among the established risk factors for stroke and vascular disease in general,^5^ primary prevention trials focusing on intensive glucose control or specific oral anti-diabetic agents showed inconsistent effects on stroke risk.^8-11^ Previous Mendelian randomization studies were underpowered to detect effects of hyperglycemia (HbA1c or fasting glucose levels) on stroke risk.^49, 50^ Here, by using data from >400,000 individuals from the UK Biobank, we were able to show that genetically elevated HbA1c levels are associated with a higher risk of ischemic stroke, thus suggesting that preventive strategies focusing on long-term HbA1c-lowering will result in risk reductions for ischemic stroke. The lack of significant effects in previous trials might relate to insufficient power due to the low number of incident stroke events, short follow-up periods, and differences in the efficacy profiles of the individual treatments.^51^

We found the effects of genetic predisposition to T2D and hyperglycemia to be specific for large artery and small vessel stroke. In accordance with these results, we found genetic predisposition to T2D to be associated with carotid plaque, an atherosclerotic phenotype, and fractional anisotropy, a marker of WM integrity associated with small vessel disease. Thus, our findings provide evidence for a causal involvement of T2D and hyperglycemia in both large artery atherosclerosis and cerebral small vessel disease. On the contrary, we found no significant effects of T2D or other diabetic traits on cardioembolic stroke. Differences in the magnitude of the effects between stroke subtypes might in part explain the heterogeneity in the effects of glucose-lowering treatments across previous clinical trials.^51^ On the basis of our findings, future trials testing glucose-lowering approaches should account for stroke subtypes.

As another major finding, we show that genetic predisposition to insulin resistance and β-cell dysfunction influences the risk of stroke independently of hyperglycemia. This could have clinical implications for oral anti-diabetic medications. While all anti-diabetic agents lower glucose levels, some drug classes primarily target insulin sensitivity whereas others primarily target β-cell function.^12, 13^ Our findings indicate that the mechanism of action might have relevance for stroke prevention on top of the glucose-lowering effects. This could further provide another layer of explanation for the heterogeneous effects of existing clinical trials testing different anti-diabetic medications for stroke prevention.^52^

Interestingly, we also found differential effects of genetic predispostion to insulin resistance and β-cell dysfunction on risk of stroke subtypes and related phenotypes. Genetic predisposition to insulin resistance showed effects on both large artery and small vessel stroke indicating more widespread effects on cerebral circulation similar to those of hyperglycemia. In contrast, genetic predisposition to β-cell dysfunction was associated with small vessel stroke, intracereral hemorrhage, and brain atrophy, which are all manifestations of cerebral small vessel disease.^53^ Given the lack of specific preventive strategies for cerebral small vessel disease, our MR results are of great interest and suggest that strategies targeting β-cell dysfunction should be explored in future studies.

Our study has several methodological strengths. The large sample size (898,130 individuals for diabetic traits and up to 514,791 individuals for stroke) and nature of our datasets provided the power to detect differential effects of diabetes on etiological stroke subtypes and to perform multiple sensitivity analyses for testing the validity of the MR assumptions, thus minimizing the possibility of biased results. While the genetic determinants of HbA1c might influence its levels via both erythrocyte and glycemic biology, we provided support for the latter, as the effects were stronger when focusing on variants not associated with erythrocyte traits. Incorporating insulin resistance and β-cell dysfunction on top of hyperglycemia in the analyses offered deeper insights into the pathophysiological mechanisms linking diabetes with the different stroke subtypes. Finally, the exploration of additional cerebrovascular disease traits enabled us to triangulate our findings for stroke subtypes by showing similar associations for etiologically related phenotypes.

Our study also has limitations. First, by design MR examines the effects of lifetime exposure to the traits of interest, which might differ from the effects of clinical interventions (e.g. glucose-lowering approaches) applied for shorter time periods later in life. Second, it was only possible to use proxies for the genetic predisposition to insulin resistance and β-cell dysfunction. However, available GWASs for biomarkers more closely related to these phenotypes (e.g. the homeostatic model assessment indexes for insulin resistance and β-cell dysfunction) were underpowered, thus precluding the identification of valid genetic instruments.^54^ Third, the MR analyses for insulin resistance were weighted based on the effects of the genetic variants on fasting insulin adjusting for BMI and the analyses for β-cell dysfunction based on the effects of the variants on fasting pro-insulin adjusting for fasting insulin. These adjustments in the original GWASs might increase the risk for collider bias in MR analyses,^55^ which should be considered when interpreting our findings. Fourth, despite using the largest currently available GWAS datasets, our analyses for some of the etiologically related traits remain underpowered. Finally, our analyses were primarily based on datasets involving individuals of European ancestry and might thus not be applicable to other ethnicities.

In conclusion, our results suggest causal associations of T2D and hyperglycemia with a higher risk for ischemic stroke, particularly large artery and small vessel stroke. Against findings from secondary analyses of clinical trials, our results support that therapeutic approaches aimed at lowering HbA1c have the potential to decrease the risk of ischemic stroke. The effects of genetically determined insulin resistance and β-cell dysfunction on large artery and small vessel stroke observed here might have implications for anti-diabetic treatments specifically targeting these mechanisms.

## Data Availability

The genetic instruments used for every exposure are made available as Supplementary material. The effects of the genetic instruments on the outcomes under study come from publicly available sources that are detailed in the Methods and the Acknowledgements sections

## Acknowledgements

This research has been conducted using the UK Biobank Resource (UK Biobank application 2532). We acknowledge the contributions by the DIAGRAM Consortium, the MAGIC Consortium, the MEGASTROKE Consortium, the ISGC Consortium, and the CHARGE Consortium for making their data publicly available. MEGASTROKE has received funding from the sources detailed at http://www.megastroke.org/acknowledgments.html.

## Author contributions

MKG, EH, RM, HSM, and MD conceived and designed the study. MKG, EH, and RM analysed the data. All authors participated in interpretation of results. MKG and MD drafted the manuscript. All authors critically revised the manuscript for intellectual content. All authors approved the submitted version and are accountable for the integrity of this work.

